# Value CMR: Towards a Comprehensive, Rapid, Cost-Effective Cardiovascular Magnetic Resonance Imaging

**DOI:** 10.1101/2020.10.09.20204305

**Authors:** El-Sayed H. Ibrahim, Luba Frank, Dhiraj Baruah, Jason C. Rubenstein, V. Emre Arpinar, Andrew S. Nencka, Kevin M. Koch, L Tugan Muftuler, Orhan Unal, Jadranka Stojanovska, John Charlson, Elizabeth M. Gore, Carmen Bergom

## Abstract

CMR is considered the gold standard for measuring heart function, including cardiac volumes and mass. Further, in a single CMR exam, information about cardiac function, structure, tissue composition, and blood flow could be obtained. Nevertheless, CMR is underutilized due to long scanning times, the need for multiple breath-holds, use of a contrast agent, and relatively higher cost compared to echocardiography.

In this study, we propose a rapid CMR exam based on recent developments in imaging sequences. The proposed exam is both rapid and provides comprehensive cardiovascular information without the need for a contrast agent or multiple breath-holds. The developed exam includes advanced sequences for evaluating global and regional cardiac functions, myocardial tissue characterization, and flow hemodynamics in the heart, valves, and large vessels. Time-consuming conventional sequences have been replaced by advanced sequences, which resulted in reducing scan time from > 1 hour with conventional CMR exam to <20 minutes with the proposed rapid CMR exam. Specifically, conventional two-dimensional (2D) cine and phase-contrast (PC) sequences have been replaced by optimized three-dimensional (3D)-cine and four-dimensional (4D)-flow sequences, respectively. Compared to 2D cine imaging that requires 12-16 separate breath-holds, the implemented 3D-cine sequence allows for whole heart coverage in 1-2 breath-holds; thus, reducing scan time by 80-90%. Similarly, compared to 2D PC flow imaging that requires multiple breath-holds and the presence of an experienced cardiac operator for precise prescription of the imaging planes, the implemented 4D-flow sequence allows for whole-chest coverage in ∼10-minute, free-breathing acquisition without the need for a navigator echo, which makes scan time independent of the patient’s breathing pattern. Furthermore, conventional myocardial tagging has been replaced by the fast strain-encoding (SENC) sequence, which reduces scan time from one slice per breath-hold to only one slice per heartbeat (∼1 second). Finally, T1 and T2 mapping sequences are included in this proposed exam, which allow for myocardial tissue characterization without the need for contrast. The proposed rapid exam has been tested on volunteers and measurements showed good agreement with those from conventional sequences despite the significant reduction in scan time.

In conclusion, we propose a rapid, contrast-free, and comprehensive cardiovascular exam that does not require repeated breath-holds or a cardiac experienced operator to run the exam, which would result in improving cost effectiveness of CMR and increasing its adoption in clinical practice.

## Background

CMR is considered the gold standard for measuring heart function, including cardiac volumes and mass (1-4). Furthermore, in a single CMR exam, information about cardiac function, structure, tissue composition, and blood flow could be obtained. Compared to echocardiography, CMR has higher reproducibility, more accuracy, and is not affected by the acoustic window, geometric assumptions, or operator’s skills (5). Nevertheless, CMR is underutilized due to long scanning times, the need for multiple breath-holds, the use of a contrast agent, and relatively higher cost compared to echocardiography (6).

In this study, we propose a rapid CMR exam based on recent developments in imaging sequences. The proposed exam is both rapid and provides comprehensive cardiovascular information without the need for contrast or multiple breath-holds, which makes it cost-effective and allows for routine adoption in clinical practice. The proposed exam includes advanced sequences for evaluating global and regional cardiac functions, myocardial tissue characterization, as well as flow hemodynamics in the heart, valves, and large vessels. The information from different sequences complement each other to provide a complete picture about heart health. Time-consuming conventional sequences are replaced in the proposed exam by advanced sequences that are mostly free-breathing or real-time and do not require breath-holding, which resulted in total scan time of less than 20 minutes, versus more than 1 hour for conventional CMR.

Cine imaging is an essential part of almost every CMR exam, which is used to evaluate heart function based on basic cardiovascular parameters such as volumes, mass, and ejection fraction (EF) (3). Nevertheless, conventional cine imaging requires long scan times due to the need to acquire a stack of short-axis (SAX) slices (typically 12-16 slices) that covers the whole heart, in addition to other long-axis (LAX) slices. As cine slices are typically acquired one slice per breath-hold, acquiring all cine images in this conventional fashion consumes a considerable amount of scan time. In the proposed rapid exam, we replaced conventional breath-holding 2D cine imaging with a single 3D-cine scan (7, 8) based on recent developments in acquisition and reconstruction techniques (9-12), which require only 1-2 breath-holds and result in 80-90% savings in scan time compared to 2D acquisition. We further evaluated the effects of different imaging acquisition strategies on image quality and resulting clinical measurements in volunteer scans.

CMR blood flow imaging is essential in certain applications, e.g. in valvular and congenital heart diseases. It also provides important information about hemodynamic patterns, diastolic function, and vessel wall stiffness in different cardiovascular diseases. Typically, 2D phase-contrast (PC) sequences are used for flow imaging, where each slice is imaged in a separate breath-hold, which results in long scan time with the possibility of scan repetitions to adjust slice positioning as accurate plane prescription is necessary for quantitative flow analysis. In the proposed exam, 2D PC flow imaging was replaced by an advanced 4D-flow imaging (13, 14) based on recent developments in acquisition and reconstruction techniques (9, 12), which allows for free-breathing acquisition, whole-chest coverage, and inline image reconstruction in ∼10 minutes, independent of the patient’s breathing pattern. This approach significantly reduces scan time and allows for measuring flow and other derivative parameters at any location in the acquired volume without the need to repeat scans. In this study, we compared flow measurements from 4D-flow acquisition to those from conventional 2D PC flow imaging at different locations.

Measuring regional cardiac function parameters, e.g. myocardial strain, showed to be valuable for early detection of subclinical cardiac dysfunction before global function, e.g. EF, is affected (15, 16). CMR tagging is typically used for strain imaging (17, 18). Nevertheless, the tagging sequence requires a separate breath-hold for each acquired slice, which makes it time consuming. Further, the resulting tagged images have limited spatial resolution, depending on tags separation, and requires specialized analysis software. In the proposed exam, CMR tagging was replaced by fast strain-encoding (SENC) imaging (19, 20), which has been previously validated against conventional tagging in several studies (21). Compared to CMR tagging, SENC imaging has the following advantages: 1) real-time (one slice per heartbeat) acquisition; 2) high spatial resolution on the pixel level; 3) instant, simple post-processing that results in color-coded strain maps. Replacing tagging with SENC results in significant saving in scan time.

Finally, the proposed exam includes T1 and T2 mapping (22, 23) sequences for myocardial tissue characterization. The advantages of these techniques are that they provide quantitative measurements of myocardial T1 and T2 relaxation parameters without the need for a contrast agent. Elevated T1 and T2 values have been previously shown to be associated with the presence of tissue fibrosis and edema, respectively (22, 24).

## Methods

Human studies were conducted with approval of our Institutional Review Board. Two healthy subjects were scanned using both the proposed rapid exam and conventional CMR sequences on a GE 3T Premier MRI scanner (GE Healthcare, Waukesha, WI) equipped with 32-channel phased-array coil. The proposed rapid CMR exam included the following sequences: 3D-cine (full heart coverage in 1-2 breath-holds), navigator-echo-free 4D-flow (whole-chest coverage in ∼10 minutes of free-breathing), SENC (3 SAX and 3 LAX slices obtained in 6 heartbeats without breathing holding), and T1 (Modified Look-Locker (MOLLI)) (25) and T2 (multi-echo fast spin echo (MEFSE)) mappings, with same coverage as in SENC imaging.

The implemented 3D cine imaging technique included a number of acquisition and reconstruction developments, including variable-density k-t sampling (12) and kat ARC acceleration (Autocalibrating Reconstruction for Cartesian Sampling with k-& Adaptive-t-space data synthesis; a spatiotemporal correlation-based auto-calibrating parallel imaging method) (9-11). The technique allows for achieving an acceleration factor of 9 and whole-heart imaging in 1-2 breath-holds. The following whole-heart coverage acquisition strategies were implemented: 1) CONV: conventional 2D cine acquisition with SAX slices acquired in separate breath-holds; 2) 2-SAX: two overlapping 3D-cine slabs acquired in the SAX direction in two breath-holds; 3) 1-SAX: one 3D-cine thick slab acquired in the SAX direction in a single breath-hold; 4) 2-AXIAL: two overlapping 3D-cine slabs acquired in the axial direction (with almost isotropic resolution) in two breath-holds; and 5) 1-AXIAL: one 3D-cine thick slab acquired in the axial direction (with almost isotropic resolution) in a single breath-hold. The imaging parameters for 2D cine imaging were: repetition time (TR) = 3.6 ms, echo time (TE) = 1.3 ms, flip angle = 55°, views per segment = 14. The imaging parameters for 3D-cine sequence were: TR = 3 ms, TE = 1.2 ms, flip angle = 40°, views per segment = 20. Both 2D cine and 3D-cine sequences were obtained with Fast Imaging Employing Steady-state Acquisition (FIESTA) acquisition, number of averages = 1, and readout bandwidth = 488 Hz/pixel. Additional information about spatial and temporal resolutions of different acquisitions is shown in **Table 1**.

**Table 1.** Characteristics of different cine acquisition strategies

| Method | # Stacks<br>x locations<br>(x overlaps) | Spatial<br>resolution<br>(mm) | Temporal<br>resolution<br>(ms) | Myocardium<br>SNR | Blood-to-myocardium<br>CNR | Total<br>scan<br>time (s) | %<br>Time<br>saving |
| --- | --- | --- | --- | --- | --- | --- | --- |
| **CONV** | 16 2D slices | 1.8x1.8x8 | 27 | 28 | 73 | 270 | -- |
| **2-SAX** | 2 x 14 x 4 | 1.8x1.8x7 | 40 | 45 | 81 | 48 | 82% |
| **1-SAX** | 1 x 20 | 1.8x1.8x7 | 40 | 10 | 30 | 20 | 93% |
| **2-AXIAL** | 2 x 34 x 8 | 2.5x2.5x2.8 | 40 | 18 | 18 | 48 | 82% |
| **1-AXIAL** | 1 x 46 | 2.5x2.5x2.8 | 40 | 20 | 15 | 21 | 92% |

CONV: conventional 2D cine acquisition; 2-SAX: 3D-cine acquisition of two overlapping slabs in the short-axis (SAX) direction; 1-SAX: 3D-cine acquisition of one thick slab in the SAX direction; 2-AXIAL: 3D-cine acquisition of two overlapping slabs in the axial direction with semi-isotropic resolution; 1-AXIAL: 3D-cine acquisition of one thick slab in the axial direction. Shown scan times include recovery time in-between repeated breath-holds. CNR, contrast-to-noise ratio; SNR, signal-to-noise ratio.

Myocardium signal-to-noise ratio (SNR) was measured as the ratio between mean myocardial signal intensity and background standard deviation (SD). Blood-to-myocardium contrast-to-noise ratio (CNR) was measured as the ratio between the mean blood-to-myocardial signal intensity difference and background SD. The cine images were processed using the cvi^42^ software (Circle Cardiovascular Imaging, Calgary, Canada). The AXIAL datasets were reformatted to generate a stack of SAX and LAX slices. The SAX and LAX images from different acquisition scenarios were semi-automatically processed to measure left ventricular (LV) volumes, mass, EF, and wall thickness of different heart segments based on the American Heart Association (AHA) 17-segment model.

The implemented 4D-flow imaging technique included a number of acquisition and reconstruction developments, including variable-density k-t sampling (increasing signal averaging at the k-space center) for acquisition-efficient motion suppression (12), kat ARC acceleration (9), and overlapping multi-slab acquisition for inflow blood signal enhancement without using a contrast agent. The technique allows for achieving an acceleration factor of 8, which allows for navigator-echo-free, whole-chest coverage in ∼10 minutes. The 4D-flow imaging parameters were as follows: 3D time-resolved gradient echo sequence, 3 slabs with 32 location per slab and 5 overlapping locations between adjacent slabs, matrix = 180×180, field of view (FOV) = 360×360 mm^2^, spatial resolution = 2×2×2.4 mm^3^, views per segment = 4, number of heart phases = 20, flip angle = 8°, readout bandwidth = 488 Hz/pixel, acceleration factor = 8, velocity-encoding (venc) = 160 cm/s, and number of reconstructed images ∼12,500. Conventional 2D PC flow images were acquired at seven locations (for comparison with 4D-flow): ascending aorta, proximal and distal descending aorta, main and right pulmonary arteries, and mitral and tricuspid valves. The 2D PC flow imaging parameters were as follows: gradient echo sequence, TR = 6 ms, TE = 3.8 ms, matrix = 192×160, FOV = 360×360 mm^2^, slice thickness = 6 mm, readout bandwidth = 488 Hz/pixel, flip angle = 25°, views per segment = 6, venc = 160 cm/s, number of heart phases = 30, and breath-holding time (not including recovery time in-between repeated breath-holds) = 15 s per slice.

The 2D PC and 4D-flow images were processed using the cvi^42^ software, where the 4D-flow images were analyzed to measure blood flow at the same planes acquired with the 2D PC flow imaging sequence. The 4D-flow images were analyzed on a DELL Precision 7540 mobile workstation (laptop) equipped with 8-core Xeon processor, 128 GB RAM, and NVIDIA Quadro RTX 3000 card, which resulted in streamlined analysis without noticeable time lags. The following parameters were measured at different locations: blood flow, velocity, volume, vascular pressure gradient, valvular regurgitation fraction, forward flow time, and ventricular early-to-atrial filling ratio (E/A) in the left and right ventricles.

SENC imaging is based on modulating the magnetization in the through-plane (slice selection) direction and then acquiring two sets of time-resolved images with different demodulation frequencies, from which tissue strain in the through-plane direction can be measured (19). The implemented SENC sequence included a number of techniques that allowed for reducing scan time to one slice per heartbeat, including: localized excitation, spiral readout, and interleaved image acquisition (20). Localized excitation is implemented by replacing one of the modulation hard radiofrequency (RF) pulses by a slice-selective pulse in the phase-encoding direction in order to reduce the excited FOV, and thus use a smaller acquisition FOV (and matrix) to accelerate acquisition without introducing aliasing artifacts. Interleaved spiral acquisition is implemented for data readout, where 4 interleaved spirals, each is 8ms long, are used instead of conventional cartesian readout. Finally, the two sets of demodulated images (called low-tuning (LT) and high-tuning (HT) images, which acquire data at low-frequency and high-frequency, respectively) are acquired in an interleaved fashion in a single heartbeat. The SENC imaging parameters were as follows: TR = 8 ms, TE = 4 ms, flip angle = 20°, slice thickness = 8 mm, matrix = 64×64, reduced FOV = 250×250 mm^2^, number of averages = 1, readout bandwidth = 977 Hz/pixel, temporal resolution = 32 ms. The SENC images were processed using the MyoStrain software (Myocardial Solutions, Inc, Morristown, NC) to measure circumferential (Ecc) and longitudinal (Ell) strains. Conventional tagging images were acquired for reference with the following imaging parameters: TR = 5.7 ms, TE = 3.1 ms, flip angle = 8°, slice thickness = 7 mm, matrix = 212×192, FOV = 360×360 mm^2^, number of averages = 1, readout bandwidth = 391 Hz/pixel, tag spacing = 7 mm, # heart phases = 30.

The imaging parameters of the MOLLI T1 mapping sequence were as follows: 8 images acquired using the 5(3s)3 acquisition pattern, FIESTA sequence, TR = 2.9 ms, TE = 1.3 ms, flip angle = 35°, slice thickness = 8 mm, matrix = 160×148, FOV = 360×360 mm^2^, number of averages = 1, and readout bandwidth = 977 Hz/pixel. The imaging parameters of the MEFSE T2 mapping sequence were as follows: TR = 895 ms, TE = 11-77 ms (4 echoes with 22ms increments), echo train length (ETL) = 16, flip angle = 90°, slice thickness = 8 mm, matrix = 180×180, FOV = 360×360 mm^2^, number averages = 1, readout bandwidth = 651 Hz/pixel. Both MOLLI and MEFSE images were analyzed using the cvi^42^ software to generate T1 and T2 maps, respectively.

Statistical t-test analysis was conducted to evaluate the significance of the measurements’ differences between different acquisition techniques (P < .001 considered significant). Further, Bland-Altman (26) analysis was conducted to assess inter-method variability between different flow imaging sequences.

## Results

Scanning times of the proposed rapid exam and conventional exam were less than 20 minutes and greater than 1 hour, respectively. The results from conventional 2D cine and different 3D cine acquisitions (**Figures 1-3**) showed good agreement between the images obtained in the same subject using different acquisition strategies. **Table 1** summarizes scan time, SNR, and CNR results of different cine acquisitions. The 2-SAX, 1-SAX, 2-AXIAL, and 1-AXIAL 3D cine acquisitions resulted in 82%, 93%, 82%, and 92% savings in scan time, respectively, compared to conventional 2D cine acquisition. SNR increased by 86% in the 2-SAX 3D cine acquisition compared to conventional 2D cine acquisition, while it decreased by 37.5%, 49.5%, and 29% in the 1-SAX, 2-AXIAL, and 1-AXIAL 3D cine acquisitions, respectively. CNR increased by 24% in the 2-SAX 3D cine acquisition compared to conventional 2D cine acquisition, while it decreased by 26.5%, 78%, and 77.9% in the 1-SAX, 2-AXIAL, and 1-AXIAL 3D cine acquisitions, respectively.

**Figure 1.**
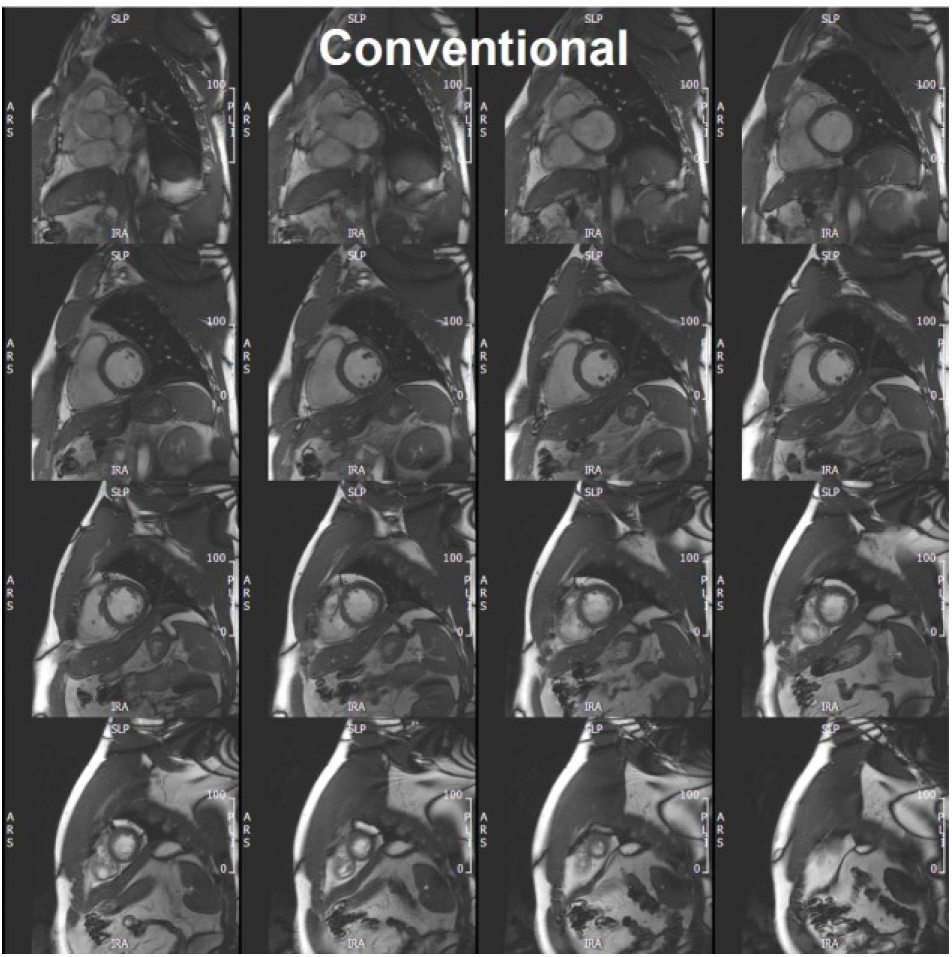
A stack of short-axis slices covering the heart from base (top left) to apex (bottom right), obtained using conventional 2D cine acquisition, where each slice is acquired in a separate breath-hold. Total scan time (including recovery time in-between consecutive breath-holds) = 270 s.

**Figure 2.**
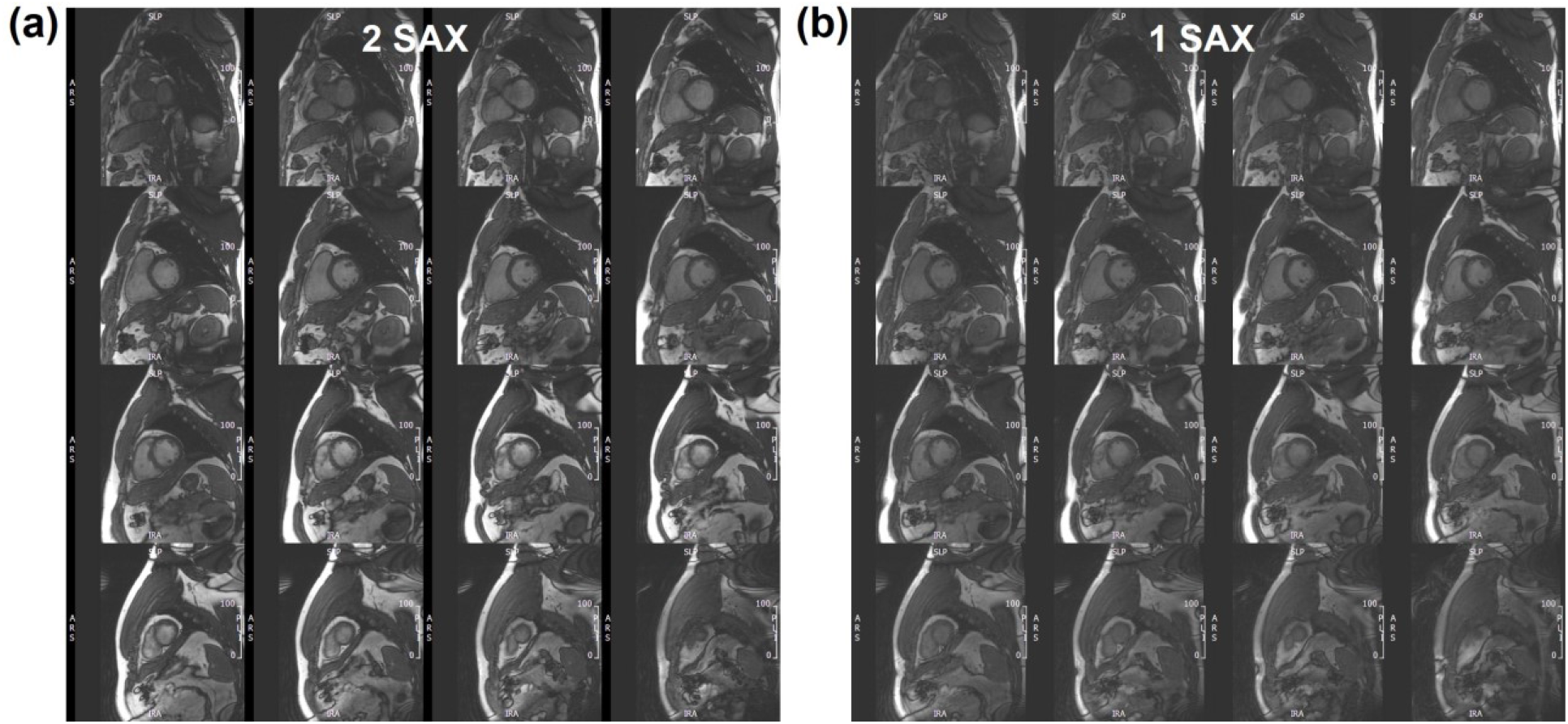
Stacks of short-axis slices covering the heart from base (top left) to apex (bottom right), obtained in the same subject as in figure 1 using accelerated 3D-cine acquisition with 2 overlapping short-axis (SAX) slabs (a) and one thicker SAX slab (b). Scan time is 48 s and 20 s for the 2-slabs and 1-slab acquisitions, respectively.

**Figure 3.**
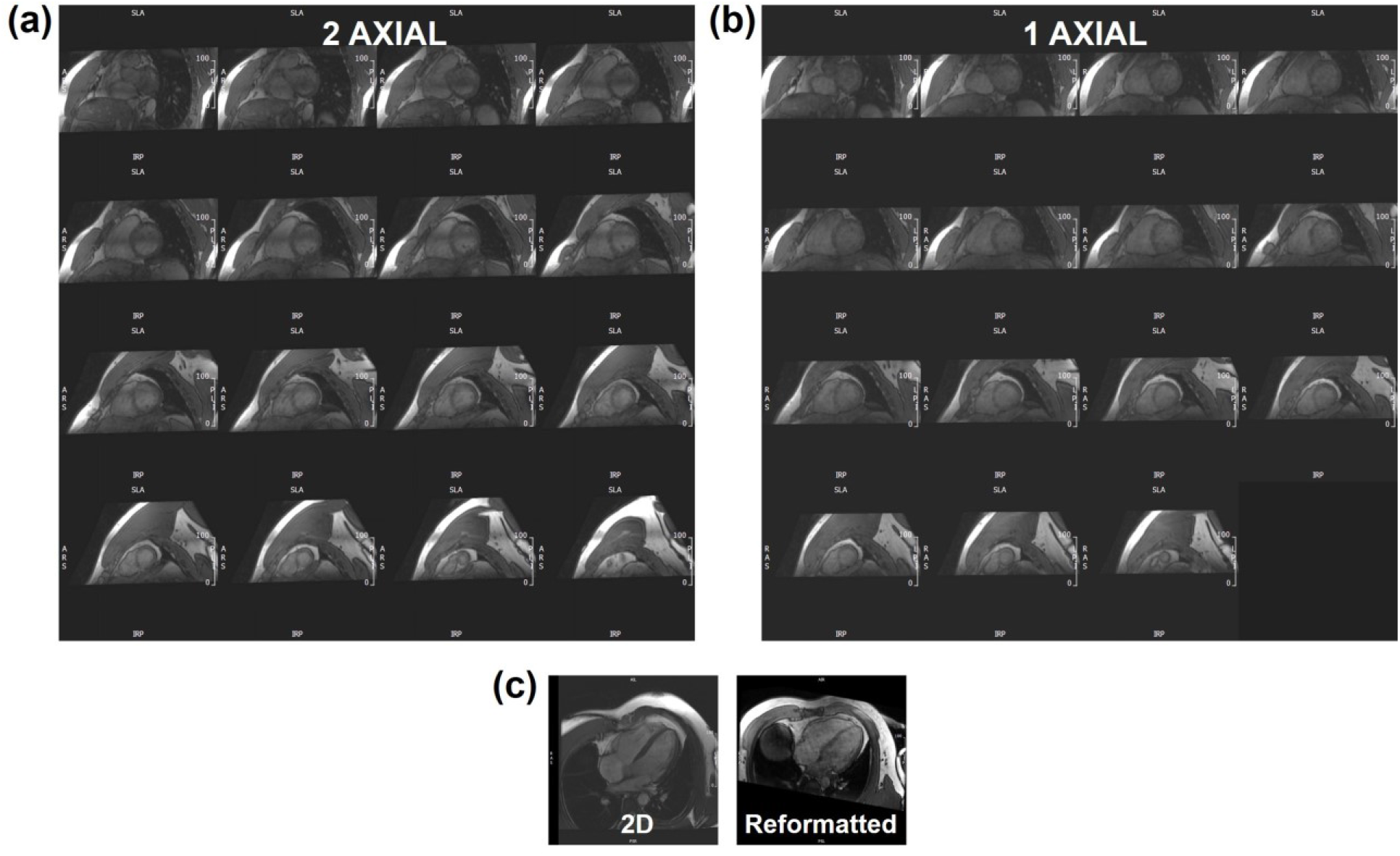
Reformatted short-axis slices, covering the heart from base (top left) to apex (bottom right), obtained in the same subject as in figures 1 and 2 using accelerated semi-isotropic 3D-cine acquisition with 2 overlapping axial slabs (a) and one thicker axial slab (b). Scan time is 48 s and 21 s for the 2-slabs and 1-slab axial acquisitions, respectively. (c) Four-chamber slice reformatted from the semi-isotropic 3D-cine acquisition (right) and a corresponding slice acquired using conventional 2D cine imaging (left).

Conventional 2D cine and AXIAL and SAX 3D cine acquisitions had similar global cardiac function results. Conventional 2D cine acquisition resulted in LV EF of 60% and 61%, and mass of 129g and 125g for the first and second subjects, respectively. Average differences in LV EF, compared to conventional 2D cine acquisition, were −8%, −4%, −7%, and −7% for the 2-SAX, 1-SAX, 2-AXIAL, and 1-AXIAL 3D cine acquisitions, respectively, while average difference in mass, compared to 2D cine acquisition, were 5g, 2g, 3g, and 1g for the 2-SAX, 1-SAX, 2-AXIAL, and 1-AXIAL 3D cine acquisitions, respectively. Average end-diastolic (end-systolic) percentage differences in segmental wall thickness, compared to 2D cine acquisition, were 1.6%, −0.7%, −5.3%, and 6.4% (−7.9%, 4.1%, −1.1%, and −8.9%) for the 2-SAX, 1-SAX, 2-AXIAL, and 1-AXIAL 3D cine acquisitions, respectively. All measurement differences were statistically insignificant.

**Figures 4** and **5** show 2D PC flow results and the corresponding 4D flow maps and flow curves at different measurement sites of large arteries (aorta and pulmonary artery) and atrioventricular valves (mitral and tricuspid), respectively. The measurements among all imaged locations and studied subjects are summarized in **Table 2**. All scans were successfully completed without motion or other imaging artifacts. Various measurements were within normal ranges reported in the literature. Among all imaged locations and studied subjects, the measurements mean ± SD were as follows from 2D PC flow (4D-Flow) imaging: max flow = 301.5±97.2 (299.6±88.7) ml/s, forward volume = 68.1±20.8 (62.5±15) ml^3^, max velocity = 55.2±26.6 (48.9±22.5) cm/s, forward flow time = 368±29.5 (341.5±25.2) ms, maximum pressure gradient = 2.9±1.9 (3.1±1.9) mmHG, mean pressure gradient = 0.6±0.3 (0.6±0.3) mmHG, and E/A ratio = 2.2±0.3 (2.1±0.5). The differences between paired measurements from both techniques were statistically insignificant. Bland-Altman plots showed that all measurement differences lying within the 2-standard deviation difference limit (except for a single point in the maximum pressure gradient plot), as shown in **Figure 6**.

**Table 2.**
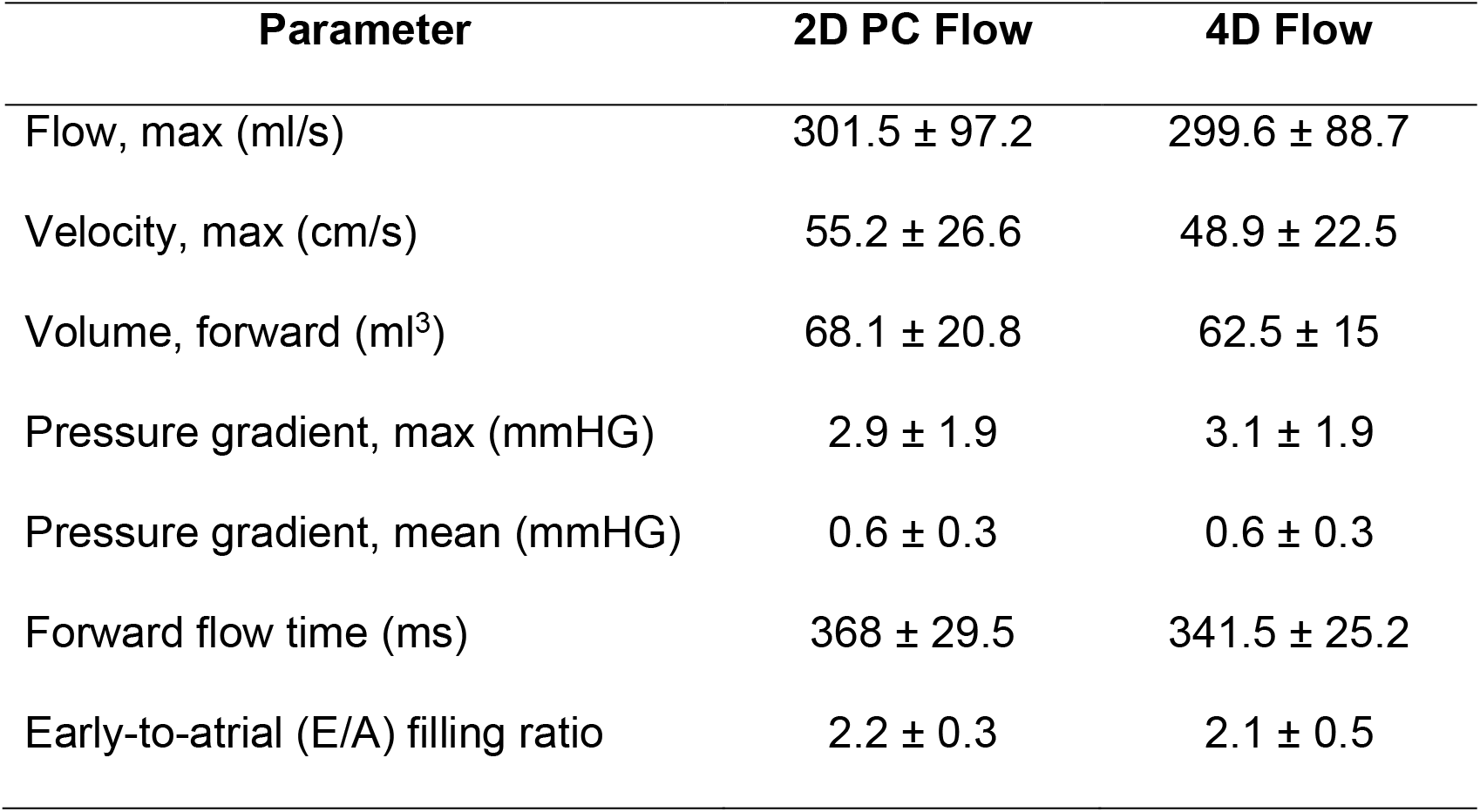
Measurements (mean±SD) of different hemodynamic parameter at the acquisition sites

**Figure 4.**
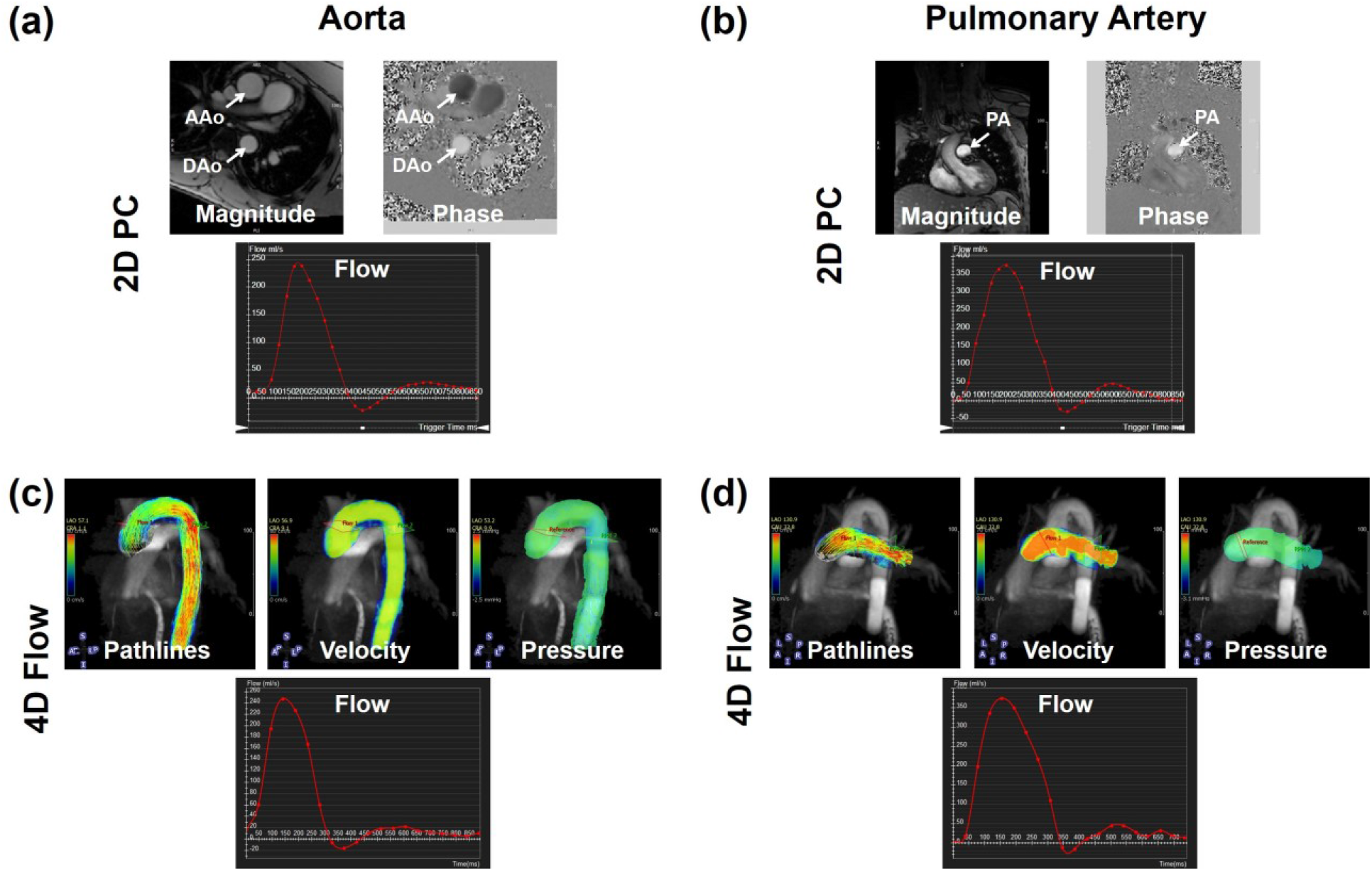
Representative 2D phase contrast (PC) images and flow curves (a, b) and the corresponding 4D-flow maps and curves (c, d) at different measurement sites of the aorta (a, c) and pulmonary artery (b, d). The images show good agreement between flow curves by the two techniques, in addition to the extra information and (pathlines, velocity, and pressure maps) provided by 4D-flow acquisition. AAo, ascending aorta; DAo, descending aorta; PA, pulmonary artery.

**Figure 5.**
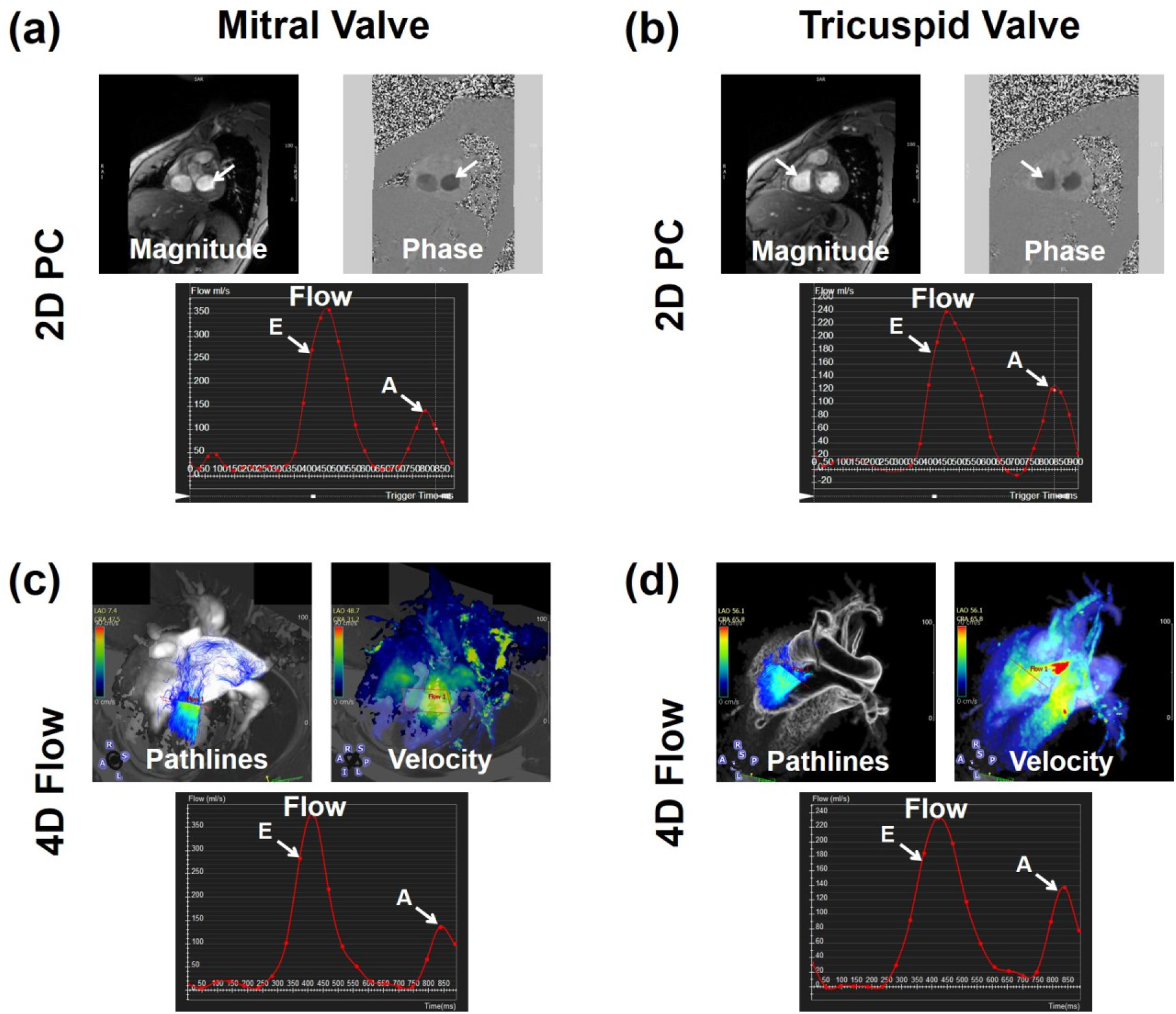
Representative 2D phase contrast (PC) images and flow curves (a, b) and the corresponding 4D-flow maps and curves (c, d) at measurement sites across the mitral valve (a, c) and the tricuspid valve (b, d). The images show good agreement between flow curves by the two techniques, in addition to the extra information (pathlines and velocity maps) provided by 4D-flow acquisition. E = early filling peak; A = atrial filling peak.

**Figure 6.**
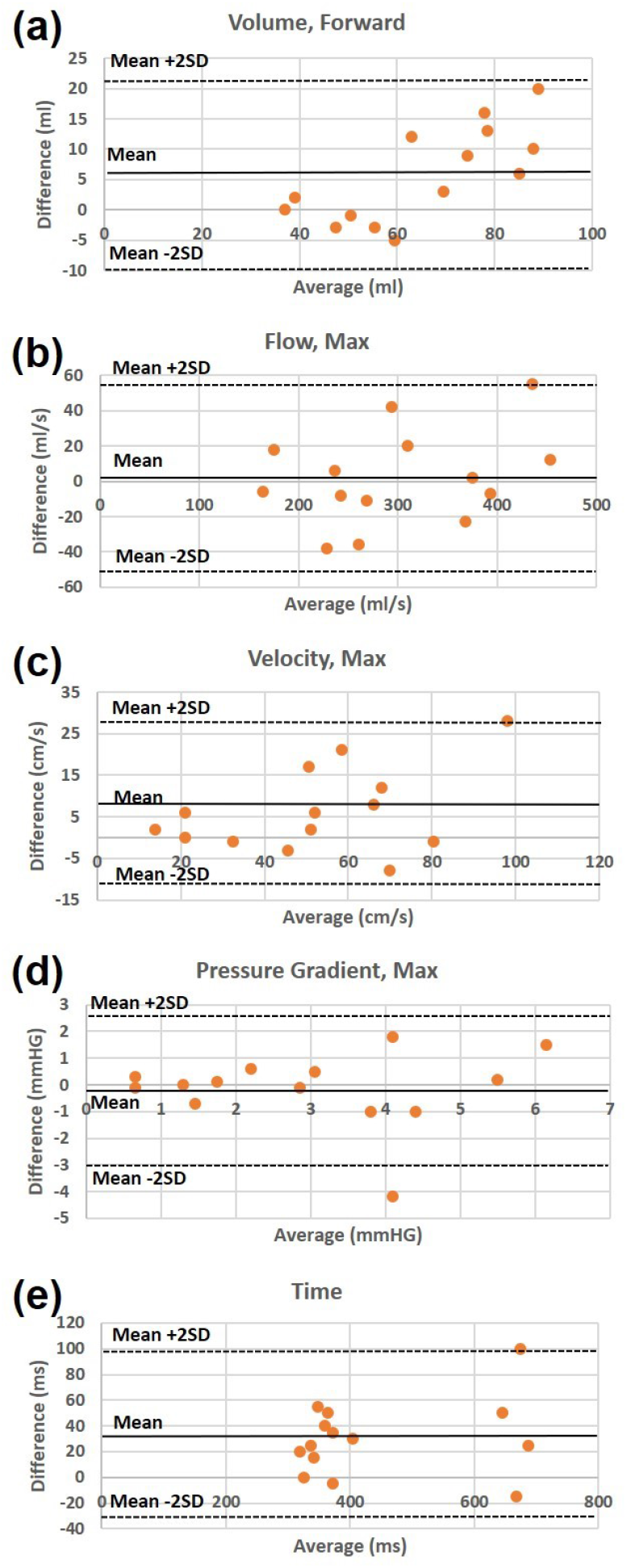
Bland-Altman plots of the differences between 2D phase contrast (PC) and 4D-flow measurements of different hemodynamic parameters acquired at different sites. (a) Forward blood volume. (b) Maximum blood flow. (c) Maximum velocity. (d) Maximum pressure gradient. (e) Forward flow time. The plots show good agreement between the two techniques as all measurement differences lie within the 2-standard deviation (SD) difference limit.

**Figure 7** shows representative SENC images and generated strain curves. The results showed normal strain values in the imaged subjects: Ecc = −23.5±2.1% and Ell = −19.7±1.9%. Corresponding tagged images are shown in the figure for reference. **Figure 8** shows resulting T1 and T2 maps, where the T1 and T2 values were within normal range in the imaged subjects: average T1 = 1233±221 ms and T2 = 48.5±1.7 ms.

**Figure 7.**
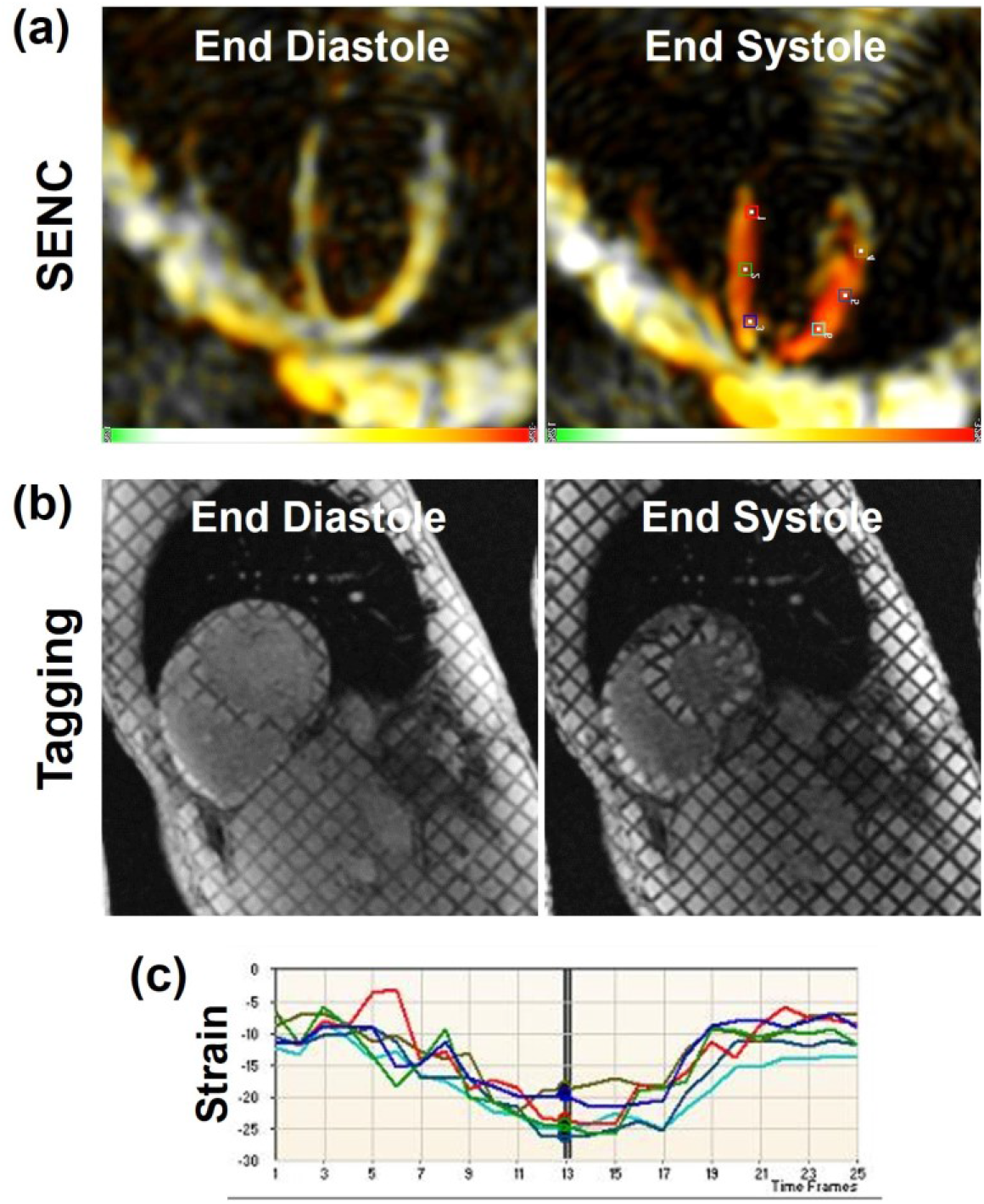
Representative strain-encoding (SENC) (a) and reference tagging (b) images acquired at both end-diastole and end-systole in the same subject. Note increased strain resolution (at the pixel level) and intuitive color-coded strain maps in SENC imaging compared to conventional tagging. (c) Circumferential strain curves generated from the SENC images at different positions shown by the markers in (a).

**Figure 8.**
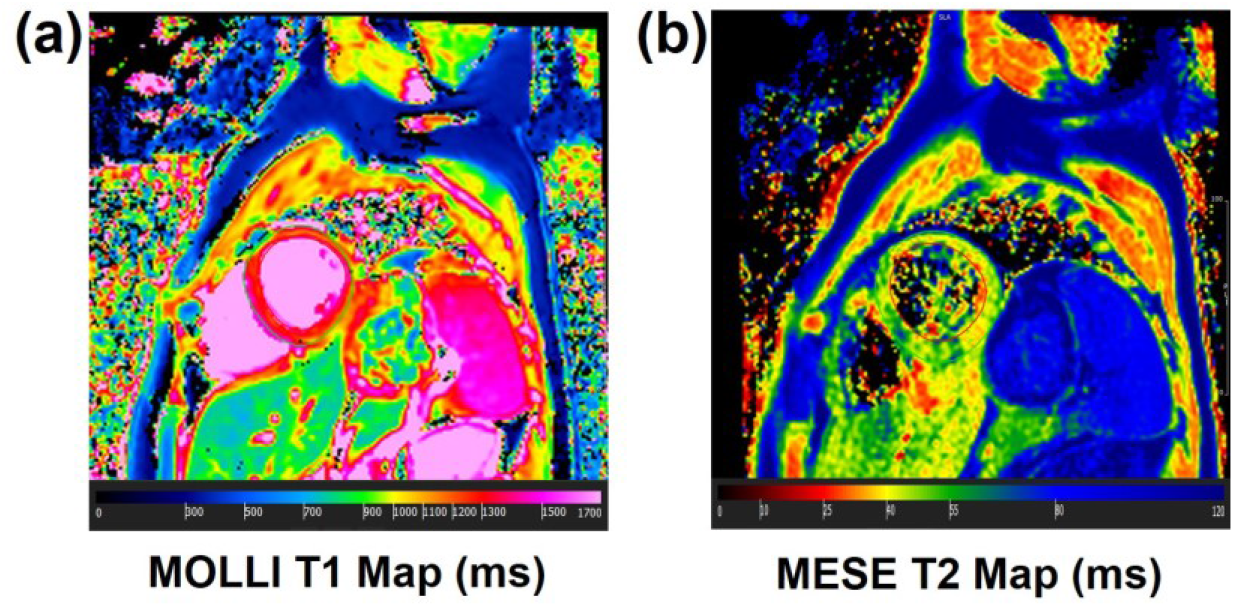
Modified Look-Locker Inversion recovery (MOLLI) T1 map (a) and multi-echo fast spin echo (MEFSE) T2 map (b) acquired in a healthy volunteer. Myocardial T1 and T2 values lie within normal range as shown by the color bars.

## Discussion

The proposed rapid CMR exam lasts for less than 20 minutes, does not require contrast, and results in comprehensive and accurate measurements of global and regional cardiac functions, hemodynamic parameters in the heart and large arteries, and T1 and T2 maps. The proposed exam makes the scan tolerable by sick patients and cost-effective and valuable from a clinical perspective.

The goal of this study was to evaluate the feasibility of the proposed rapid, comprehensive CMR exam based on advanced imaging techniques. Despite the limited number of imaged subjects, preliminary results from this study show the capability of advanced 3D-cine imaging with the proposed acquisition strategies of significantly reducing cine scan time by 80-90%. Despite encountered reduction in spatial/temporal resolutions, SNR, and CNR, sufficient image quality was maintained such that the resulting clinical measurements were not significantly different from those obtained using conventional 2D cine acquisition. The single breath-hold 3D-cine acquisition, which is more feasible for smaller patients, further reduces total scan time and avoids slice mis-registration errors. Nevertheless, in patients with limited breath-holding capability, the single breath-hold can be split in 2 shorter breath-holds for 2 slabs acquisition. AXIAL 3D acquisitions save more scan time, as no additional LAX slices need to be acquired (they are rather reformatted from the acquired semi-isotropic 3D dataset) and no experience is needed for slice prescription (the operator only needs to place an axial box covering the heart). Nevertheless, it should be noted that AXIAL 3D acquisition has reduced blood-to-myocardium CNR compared to SAX 3D acquisition due to the time-of-flight effect in the latter when fresh unsaturated blood spins flow perpendicular to and into the SAX slab, resulting in brighter blood signal and therefore higher blood-to-myocardium CNR. Furthermore, switching from two (overlapping) thinner slabs acquisition to one thicker slab acquisition causes more reduction in CNR due to the reduced time-of-flight effect as blood travels a larger distance through the thicker slab, and thus becomes more saturated and has lower signal. It should be noted that compressed-sensing-based single-heartbeat cine imaging has been recently introduced by some MRI vendors (27), which could have been used in place of conventional 2D segmented cine acquisition with significant saving in cine scan time as well.

The implemented accelerated 4D-flow technique allows for acquiring detailed whole-chest hemodynamic information in ∼10 minutes. Measurements from accelerated 4D-flow were in good agreement with conventional 2D PC flow imaging. As motion correction is not based on navigator-echo tracking (12), the imaging time is known in advance and is not dependent on the patient’s breathing pattern, which allows for efficient use of the scan time and the capability of adjusting the imaging parameters to reduce the 4D-flow scan time to a specified time duration. 4D-flow imaging has several advantages compared to 2D PC flow imaging. Besides reduction in total scan time and alleviating the need for repeated breath-holds and possible scan repetitions, prescription of the 4D-flow scan is quite simple (only placing a box around the heart), and therefore, compared to 2D PC flow imaging, no cardiac-specialist scanner operator is needed to run the 4D-flow sequence. Further, post-processing and image analysis can be done at any plane of interest and is not restricted to the slices acquired during the CMR exam as in 2D PC flow imaging. Besides its short acquisition time, the short reconstruction time (∼12,500 images are reconstructed inline within a couple of minutes) makes the scan feasible for clinical adoption as other sequences can be run on the scanner after the 4D-flow data acquisition without congesting the reconstruction pipeline. Finally, with recently available computers with large computational powers and cloud computing options, analysis of 4D-flow dataset can be done in realtime without experiencing time lags, as done in this study.

One key advantage of the proposed rapid exam is using SENC for fast strain measurements in only 6 seconds. A large body of literature showed the value of myocardial strain analysis for early detection of subclinical cardiac dysfunction in different cardiovascular diseases before global heart function is affected or symptoms of cardiovascular diseases are observed (28-30), especially in coronary artery disease (31) and heart failure (32), which would allow for early intervention and better outcomes. Further, besides providing strain measurements, SENC showed the capability of producing reproducible global function measurements that are in close agreement with those obtained from cine images (33-35). Therefore, for further reduction of scan time, more SENC images (a total of 6 SAX and 6 LAX slices) could be acquired to provide volumes and mass measurements in lieu of using cine imaging.

It should be noted that other fast and valuable CMR sequences have been recently developed, which could be added to the proposed rapid CMR exam to provide more information with only slight increases in scan time. One example of these sequences is myocardial blood oxygenation level dependent (BOLD)-based imaging (36, 37), which can provide useful information about perfusion and myocardial vasodilation after hyperventilation maneuvers and without the need for exogeneous contrast agent or pharmaceutical stress agents (36). Therefore, adding the BOLD sequence to the proposed exam would add one more dimension, in addition to T1 and T2 mappings, for tissue characterization.

One limitation of the proposed rapid exam is that it does not include perfusion or late gadolinium enhancement (LGE) sequences for ischemia and viability imaging, respectively. Nevertheless, both perfusion and LGE imaging require contrast administration and increase the scan time, especially with the delay time post contrast injection and before LGE imaging. Furthermore, with recent concerns about gadolinium side effects and accumulation in different body organs, more efforts are being made towards more adoption of contrast-free CMR sequences in clinical practice (38). It should be noted that some non-contrast-enhanced sequences showed the capability of identifying different tissue types, which could help for tissue characterization. For example, strain imaging showed the capability of differentiating between viable and infarcted myocardium (39-41) as well as identifying ischemic defects (42, 43) with results in agreement with LGE and perfusion images, respectively. Furthermore, fibrosis and myocardial infarction have higher T1 values compared to normal myocardium (22, 24); therefore, the combination of functional and T1 and T2 mappings (23) could allow for tissue characterization without the need for gadolinium. Although in this work we showed the feasibility of the proposed rapid CMR exam and its advantages compared to conventional CMR sequences, larger studies on different patient populations would be valuable to further illustrate the clinical usefulness of the proposed exam.

## Conclusions

In conclusion, the proposed CMR exam is rapid, contrast-free, and provides comprehensive information about the cardiovascular system, including early development of subclinical cardiac dysfunction. Furthermore, as advanced free-breathing, real-time, and 3D sequences are used in the proposed rapid exam, no repeated breath-holds nor specific planes prescription is needed, which increases patient comfort and allows non-cardiac-experienced scanner operators to run the exam. These features are essential for improving cost-effectiveness of CMR and increasing its adoption in clinical practice, especially in asymptomatic patients with cardiovascular risk factors or patients at-risk of developing heart failure who cannot withstand a long CMR exam.

## Data Availability

Data available upon reasonable request from the corresponding author

